# A Lightweight, End-to-End Explainable, and Generalized attention-based graph neural network to Classify Autism Spectrum Disorder using Meta-Connectivity

**DOI:** 10.1101/2024.07.17.24310610

**Authors:** Km Bhavna, Niniva Ghosh, Romi Banerjee, Dipanjan Roy

## Abstract

Recent technological advancement in Graph Neural Networks (GNNs) have been extensively used to diagnose brain disorders such as autism (ASD), which is associated with deficits in social communication, interaction, and restricted/repetitive behaviors. However, the existing machine-learning/deep-learning (ML/DL) models suffer from low accuracy and explainability due to their internal architecture and feature extraction techniques, which also predominantly focus on node-centric features. As a result, performance is moderate on unseen data due to ignorance of edge-centric features. Here, we argue that meaningful features and information can be extracted by focusing on meta connectivity between large-scale brain networks which is an edge-centric higher order dynamic correlation in time. In the current study, we have proposed a novel explainable and generalized node-edge connectivity-based graph attention neural network(Ex-NEGAT) model to classify ASD subjects from neuro-typicals (TD) on unseen data using a node edge-centric feature set for the first time and predicted their symptom severity scores. We used ABIDE (I and II) dataset with a large sample size (Total no. of samples = 1500). The framework employs meta-connectivity derived from Theory-of-Mind (ToM), Default-mode Network (DMN), Central executive (CEN), and Salience network (SN) that measure the dynamic functional connectivity (dFC) as a flow across morphing connectivity configurations. To generalize the Ex-NEGAT model, we trained the proposed model on ABIDE I(No. of samples =840) and performed testing on the ABIDE II(no. of samples =660) dataset and achieved 88% accuracy with an F1-score of 0.89. Additionally, we identified symptom severity scores for each individual subjects using their meta-connectivity links between relevant brain networks and passing that to Connectome-based Prediction Modelling (CPM) pipeline to identify the specific large-scale brain networks whose edge connectivity contributed positively and negatively to the prediction. Our approach accurately predicted ADOS-Total, ADOS-Social, ADOS-Communication, ADOS-Module, ADOS-STEREO, and FIQ scores.

## Introduction

Autism spectrum disorder (ASD) is a pervasive neurodevelopmental disorder that impairs an individual’s social behaviour and communication abilities [1, 2]. Individuals with ASD face several challenges in their day-to-day lives and often acquire concurrent disorders such as depression, anxiety, and ADHD, which may further complicate the clinical diagnosis, particularly in younger children [3]. While no accurate treatment for autism is currently available, early detection may be advantageous in alleviating primary symptoms of autism by providing appropriate interventions [3, 2]. Functional magnetic resonance imaging (fMRI) is an effective method that has provided a deeper understanding of the pathophysiology of ASD [4]. According to previous studies [5, 6, 7] using fMRI, it has been found that autism primarily affects the abilities of social cognition, interaction, and communication abilities that are associated with alternations in the specific functional brain networks such as Theory of Mind(ToM), Default Mode Network(DMN), Central Executive Network(CEN), and Salience Network(SN).

Previous research on ASD classification and prediction has majorly employed statistical methods and models, specifically voxel, ROI, and network-level analysis, and applying statistical tests such as group-level independent t-test, to assess statistical disparities between autistic and typically developing(TD) groups [8, 9, 10]. These statistical techniques have also been used as feature selection techniques to enhance the ML/DL classification performance [11, 12, 13]. The current era, marked by the availability of big data and Artificial Intelligence (AI) frameworks, provides an unprecedented opportunity to leverage connectome-based features associated with ASD. Further, the recent availability of large neuroimaging datasets opens up excellent opportunities to develop data-driven DNN models for hypothesis-driven and exploratory studies [14]. Numerous previous studies [15, 16, 17] have also incorporated ML-based techniques for feature selection, such as the least absolute shrinkage and selection operator (LASSO) and its variations. ML/DL algorithms such as SVM, variational autoencoder, recurrent neural networks, and convolutional neural networks achieved considerable performance in dealing with computationally aided diagnosis of ASD [18, 19, 20, 21].

Beyond ML models, Deep learning models, such as Convolutional Neural Networks (CNN), are an effective and scalable approach for distinguishing between ASD and TD without requiring manual feature engineering [22, 23]. Although CNN operates well with grid-like input in Euclidean space, such as naturalistic images, it is probably unsuitable for use with non-Euclidean data, like brain imaging [24]. Neuronal activity in many distinct brain regions is known to be closely linked and exhibits higher-order correlations, involving large-scale functional brain networks during rest and task [23]. Consequently, the geometric distance between distinct brain areas in Euclidean space may not effectively reflect the functional distance. Some geometric deep learning approaches, such as graph neural networks (GNNs), may circumvent this constraint and better cope with non-Euclidean data types like graphs [25, 26, 27]. The previous studies used GNN and attention-based GNN for ASD classification in two ways: either using node-centric features [28, 29], in which functional connectivity along with phenotypic features (e.g., age, gender, and ASD scores) were used to train the model at the group level, or using graph-based features set [30, 31], in which node centralized local information (or in other words, each sample were treated as a graph) was used to train the model. This GNN architecture has performed modestly due to the following limitations. In many of the previous approaches, the models mainly focus on static or node-centric features in spatial space, so the importance of edge-centric features depicting relationships between nodes is largely ignored [28, 32, 33]. This aspect of GNN architecture, based on the features above that contributed to the heterogeneity of ASD, has not been completely uncovered. Secondly, due to the high dimensionality of the GNN features, significant symptomatic variations, and heterogeneity of data across different sites, the previous GNN models gave low reliability, robustness, and generalizability in identifying ASD. Lastly, the internal architecture of GNN models takes time to execute information from one node to another, which increases the execution time to process data or, in other words, increases resource consumption. These limitations collectively contribute to overfitting the classification model and lead to unsatisfactory identification performance on independent datasets [33]. Hence, there is a genuine need for an explainable GNN architecture that can utilize both edge- and node-centric features, along with the sufficient generalizability of the proposed model architecture.

In this work, we designed an attention-based GNN framework to identify ASD from TD and detect the potential biomarkers related to ASD. Our work here mainly focuses on two fundamental research questions that can potentially improve the classification of ASD samples on unseen datasets. How can we identify a unique spatio-temporal feature set for resting-state fMRI data that can improve the classification of ASD? Previously [18, 21], functional connectivity (time-averaged) matrices were widely used to classify ASD, which fell short at times to fully capture the complexity of brain dynamics on unseen datasets, or feature engineering was used in which the model identified features from time-series data that increased time and space complexity of the model. These motivated us to identify an improved feature set that can retain the topology information of brain images by constructing individual graph data based on both node-and-edge-centric features. Furthermore, how can we propose a better explainable and generalized GNN model architecture that can achieve ideal classification? The GNN models [25, 26, 27] generally suffer from an issue of the execution time of information from one node to another. This opens up a way to identify the modified architecture of GNNN that could reduce execution time without losing any information about node-edge connectivity, as well as capture the topology of brain networks, which could improve the performance of the model by detecting potential biomarkers for ASD.

The contribution of the current study is two-fold: a) We hypothesized that higher-order correlation capturing node and edge-centric temporal features between ToM, DMN, CEN, and SN brain networks could be potential imaging biomarkers to classify autistic from typical developing without ad hoc feature engineering. To this end, we calculated meta-connectivity matrices ToM, DMN, CEN and SN functional brain networks based on resting-state fMRI data for each individuals. b) Next, we hypothesized that the higher-order correlation could faithfully track ongoing resting brain dynamics in large-scale brain networks but also require significant computational power. Hence, we proposed a generalized Ex-NEGAT model that could reduce execution time as well as capture complex brain dynamics patterns by dividing the graphs into sub-graphs using the depth-first search (DFS) approach, in which meta-connectivity matrices were subjected as input to train the model. We trained the proposed model on ABIDE I dataset and to make model generalizable, we performed testing on ABIDE II dataset. To validate our results, we performed leave-one-out and five-fold cross-validation approaches.

Additionally, to test whether meta-connectivity could be a reliable predictive biomarker, we have performed prediction of symptom severity scores for each individual participant using a connectome-based prediction modeling (CPM)approach. To the best of our knowledge, our approach based on generalized Ex-NEGAT model surpasses previous such attempts of using GNN models in classifying ASD from TD based on the novel metaconnectivity based higher-order feature. The proposed framework reduce execution time that can lead to a reduction in computational resources, which helps in overcoming the earlier-mentioned challenges and improving existing approach based on derived attention maps from the framework to search and explain the potential image biomarkers for ASD. (refer to Figure 1).

**Figure 1:**
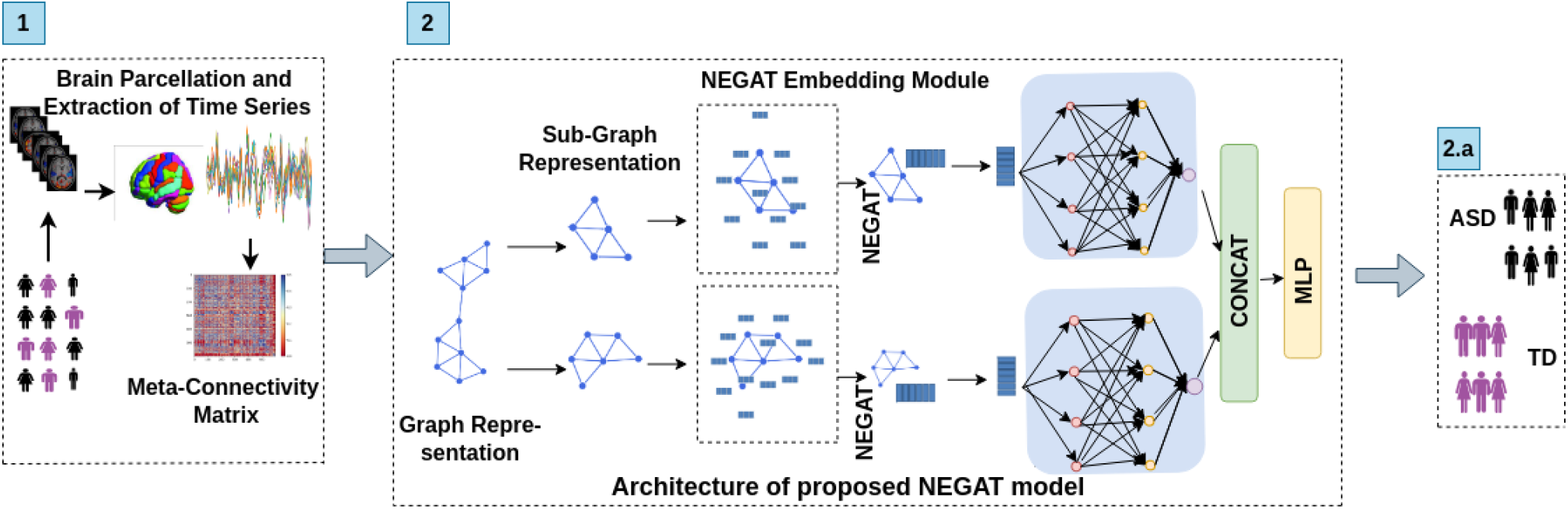
Illustrative overview of light-weight, generalized proposed Ex-NEGAT framework for classifying ASD from TD using meta-connectivity as an edge-centric feature set. Step 1 involves BOLD time-series signal extraction and calculation of meta-connectivity matrices; Step 2 shows the details of the proposed architecture NEGAT framework to perform classification.

## 2 Materials and Methods

### 2.1 fMRI Dataset and Preprocessing

In this study, we have used fMRI anatomical and resting-state functional neuroimaging data from the Autism Brain Imaging Data Exchange (ABIDE I and ABIDE II) datasets [14]. The functional scan parameters listed per site were utilized for further preprocessing. ABIDE I contains preprocessed images that were used for the study. The data acquired from ABIDE II was further preprocessed, keeping in line with the process utilized to preprocess the ABIDE I dataset. The data was preprocessed using the DPARSF (Data Processing Assistant for Resting-State fMRI) toolbox, which is compatible with MATLAB. The first 4 time points were removed to account for the time taken by the subject to settle down. The reference slice for slice timing correction was set as the slice acquired in the middle of the slice order. The Friston-24 model was employed for head motion correction, and timeframes with head motion above 0.2 mm, along with one previous and two following timeframes, were added as regressors. Nuisance regression was done using SPM apriori masks, and global signal regression was not employed as it is a contentious preprocessing measure. A bandpass filter of 0.01 to 0.1 Hz was applied, and the data was normalized using DARTEL. Smoothing was completed with a FWHM kernel size of 6 mm. Scrubbing was done for all timeframes with a head motion above 0.5 mm and one previous and two following timeframes (After preprocessing: total no. of subjects = 1500, comprised of 711 ASD and 789 TD individuals) (refer to Tables 1 and 2). The configuration for computational processing used in this study was as follows: GPUs: 16X NVIDIA Tesla V100, NVIDIA CUDA Cores: 81920, NVIDIA Tensor Cores: 10240.

**Table 1:** Demographic information of ASD and TD groups from ABIDE dataset.

| Sr. No. | Age (In Years) | Samples | IQ | ADI-R Social | ADI-R Verbal |
| --- | --- | --- | --- | --- | --- |
| ASD | Mean:13.9±6.6, S.D:6.6(range: 6 to 42) | 711 | 105±17 | 18.8±5.5 | 15.4±4.5 |
| TD | Mean:14.95±5.9, S.D:9(range: 6 to 40) | 789 | 112±14 | - | - |

**Table 2:**
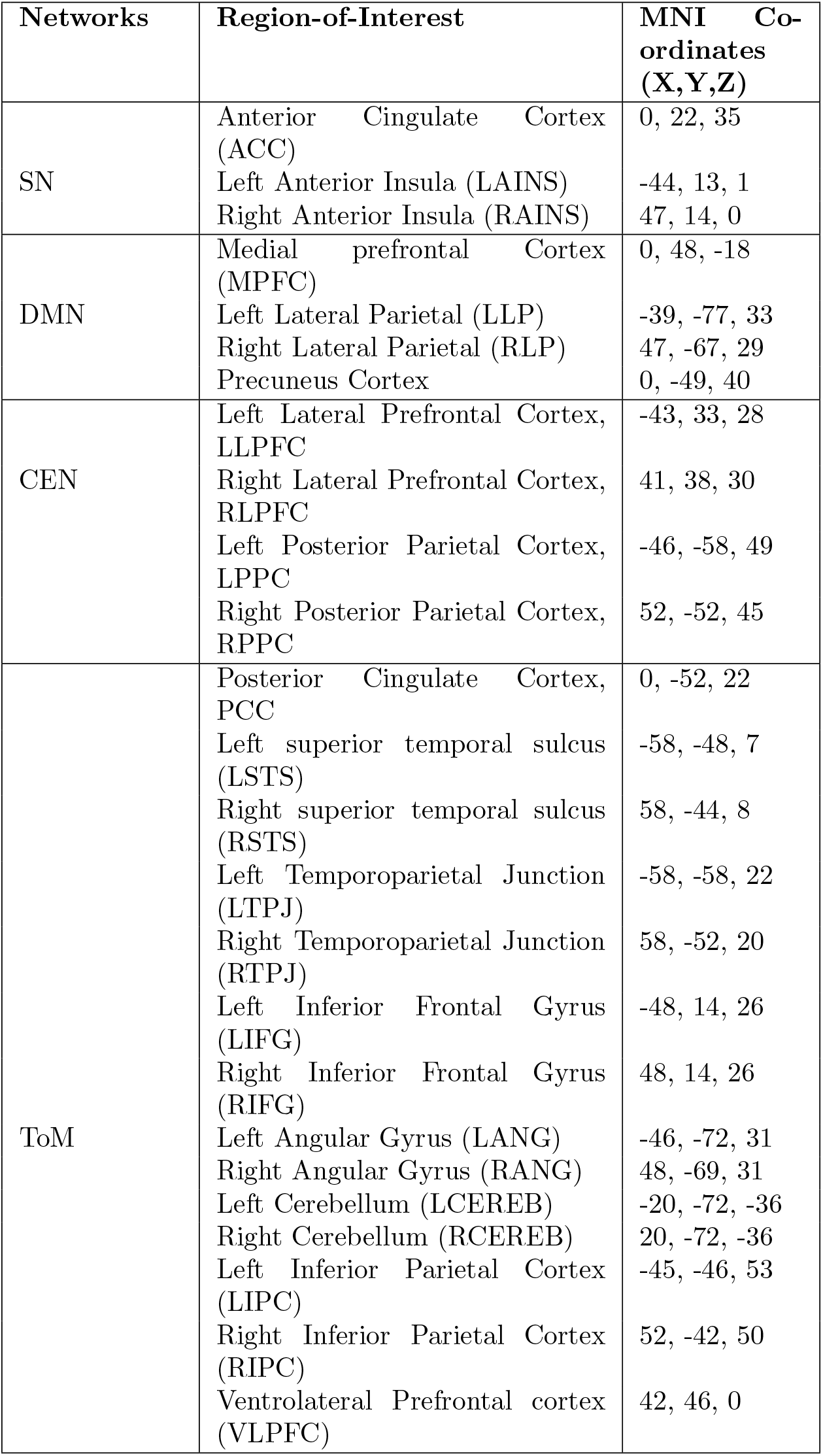
Brain region with MNI coordinates that are associated with Theory-of-Mind, Default-mode-Network, Central Executive, and Salience Networks.

| Networks | Region-of-Interest | MNI Co-ordinates (X,Y,Z) |
| --- | --- | --- |
| SN | Anterior Cingulate Cortex (ACC) | 0, 22, 35 |
|  | Left Anterior Insula (LAINS) | -44, 13, 1 |
|  | Right Anterior Insula (RAINS) | 47, 14, 0 |
| DMN | Medial prefrontal Cortex (MPFC) | 0, 48, -18 |
|  | Left Lateral Parietal (LLP) | -39, -77, 33 |
|  | Right Lateral Parietal (RLP) | 47, -67, 29 |
|  | Precuneus Cortex | 0, -49, 40 |
| CEN | Left Lateral Prefrontal Cortex, LLPFC | -43, 33, 28 |
|  | Right Lateral Prefrontal Cortex, RLPFC | 41, 38, 30 |
|  | Left Posterior Parietal Cortex, LPPC | -46, -58, 49 |
|  | Right Posterior Parietal Cortex, RPPC | 52, -52, 45 |
| ToM | Posterior Cingulate Cortex, PCC | 0, -52, 22 |
|  | Left superior temporal sulcus (LSTS) | -58, -48, 7 |
|  | Right superior temporal sulcus (RSTS) | 58, -44, 8 |
|  | Left Temporoparietal Junction (LTPJ) | -58, -58, 22 |
|  | Right Temporoparietal Junction (RTPJ) | 58, -52, 20 |
|  | Left Inferior Frontal Gyrus (LIFG) | -48, 14, 26 |
|  | Right Inferior Frontal Gyrus (RIFG) | 48, 14, 26 |
|  | Left Angular Gyrus (LANG) | -46, -72, 31 |
|  | Right Angular Gyrus (RANG) | 48, -69, 31 |
|  | Left Cerebellum (LCEREB) | -20, -72, -36 |
|  | Right Cerebellum (RCEREB) | 20, -72, -36 |
|  | Left Inferior Parietal Cortex (LIPC) | -45, -46, 53 |
|  | Right Inferior Parietal Cortex (RIPC) | 52, -42, 50 |
|  | Ventrolateral Prefrontal cortex (VLPFC) | 42, 46, 0 |

### 2.2 Calculation of Meta-Connectivity Feature-set

#### 2.2.1 Calculation of Functional Connectivity and Dynamic Functional Connectivity Stream

We extracted time-series signals from ToM, DMN, CEN, and SN brain networks to calculate meta-connectivity (refer to Table 2). Based on previous literature, we considered 25 Region-of-Interest (ROIs) [34, 35, 36, 37]. Using MNI coordinates, we created a spherical binary mask with a 10 mm radius for all selected ROIs and extracted time-series signals. We computed functional connectivity for each individual using eq (1) [38]:

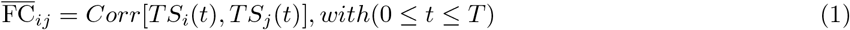

Where, *TS*_*i*_(*t*) and *TS*_*j*_(*t*) are time-series signals from *ith* ROI and *jth* ROI at time *t*, 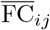 indicates average functional connectivity between ROI *i* and ROI *j*, and *t* represents specific time-point within the total time-period *T*. The functional connectivity with pair-wise Pearson correlation matrices were useful for establishing group level differences.

To track change in functional connectivity patterns over time, we calculated Dynamic Functional Connectivity (dFC) using a sliding window based approach that computed average FC matrices at different time points. According to the previous study [38], we denoted it as dFC-stream. We calculated temporal frame of dFC stream for each individual [38]:

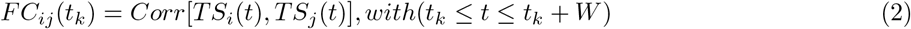

Where *t*_*k*_ indicates the *kth* temporal frame’s start time and *w* represents fixed window length. We choose *w* = 20 − 30*sec*, according to the literature [39]. Sliding step Δ*τ* is used to separate frame start time, such that *t*_*k*_ = *t*_*k*−1_ + Δ*τ* = *k*Δ*τ*.

#### 2.2.2 Meta-Connectivity Analysis

Each dFC stream was considered as a collection of time series that defines the time dependency of individual FC pairwise couplings. An *M* * *M* matrix of correlations between the time-dependent strengths of *M* = *N* (*N* − 1) FC links (*N* ^2^ pairs of regions, minus the self-loops) was produced by improving the calculation of FC matrix from regional nodes to inter-regional links [38]. The resultant Meta-Connectivity (MC) matrix expresses the inter-link covariance analogous to how the FC matrix explains the inter-node covariance (refer to Figure 2).

**Figure 2:**
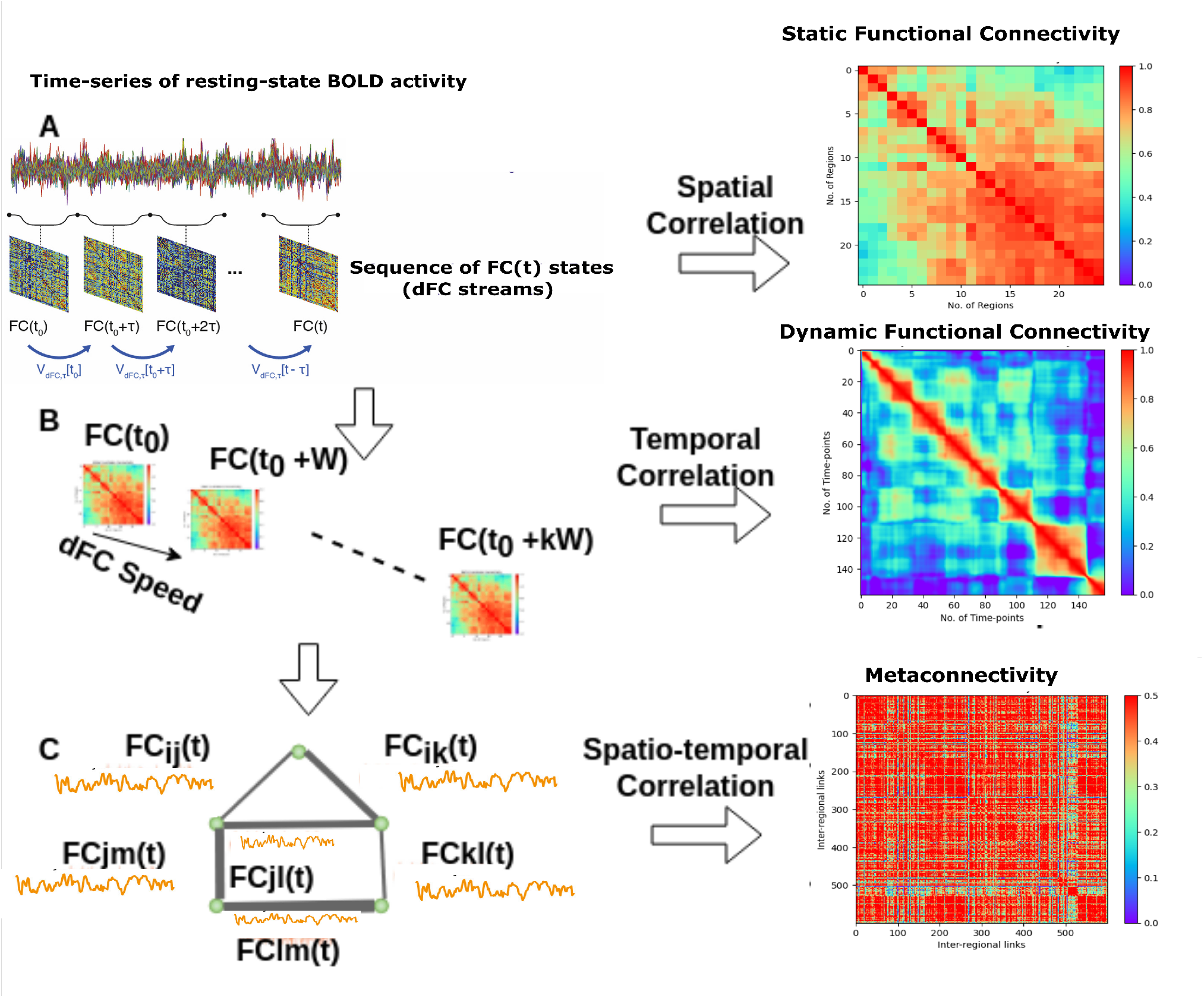
Calculation of Static, Dynamic functional connectivity, and Meta-connectivity: In the traditional approach **(A)**, neural activity is averaged over time to create a functional connectivity (FC) matrix. We calculated a dynamic FC stream using shorter sliding windows **(B)**, leading to a dynamic FC matrix. In **(C)**, we considered each FC link as a dynamic variable, giving rise to a Meta-Connectivity (MC) matrix by capturing covariances between these dynamic links.

**Figure 3:**
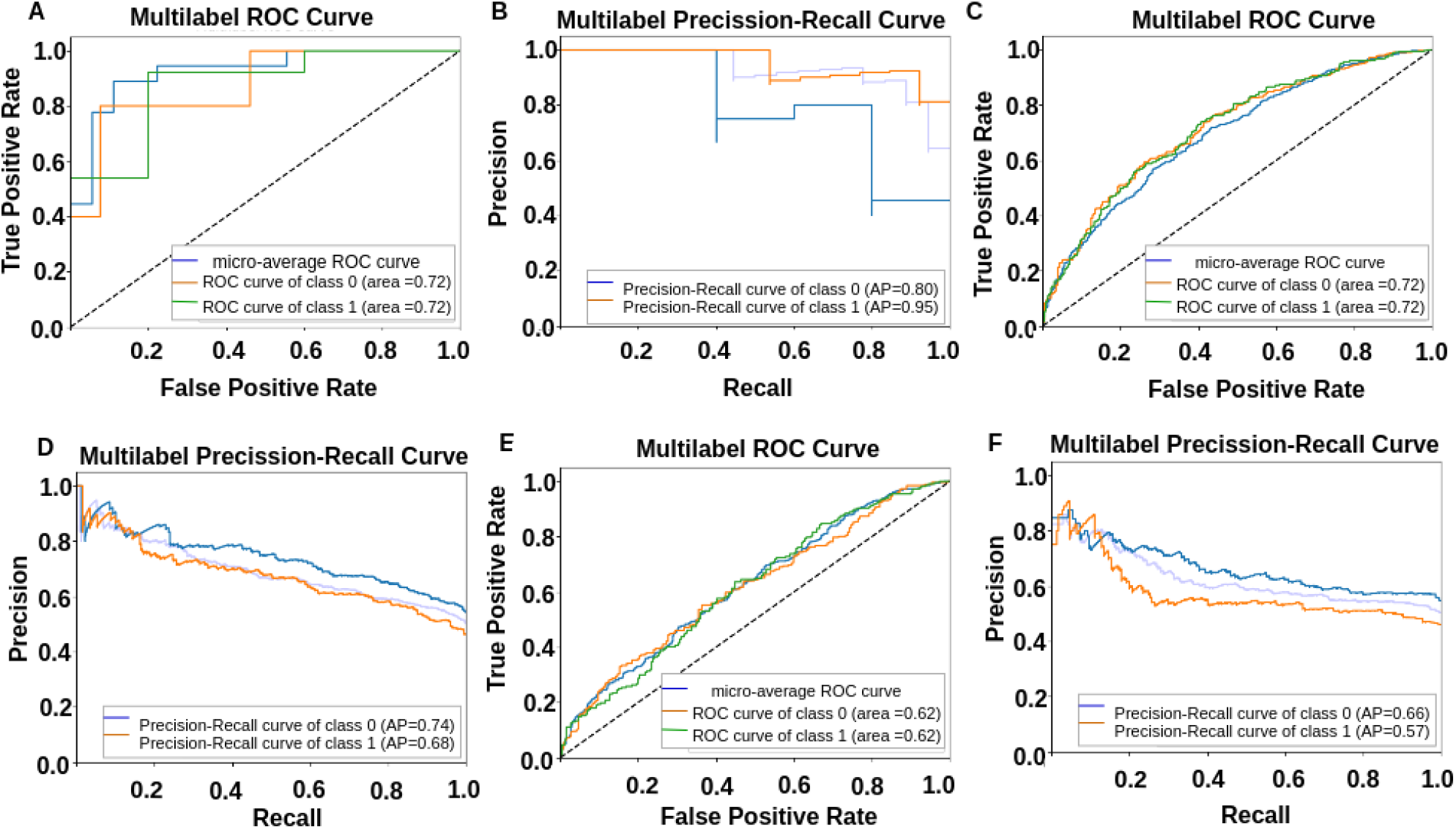
**A) and B)** are depicting classification results using meta-connectivity with window size =30 sec. **C) and D)** are depicting classification results using meta-connectivity with window size = 20 sec. Whereas **E) and F)** are depicting classification results using functional connectivity. **G) and H)** are showing classification results using Dynamic Functional Connectivity.

To calculate MC matrices, we used dFC stream and extracted *n* = *N* (*N* − 1)/2 time-series of pairwise FC coupling, in which input was given as *FC*_*ij*_(*t*) for all pair of regions i and j with *i < j* ≤ *N* using equation (3) [38]:

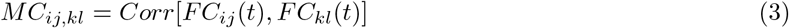

These *MC*_*ij,kl*_ entries are computed into a matrix format, making it possible to identify the pair of links contributed in each meta-link directly. Consequently, MC matrices had a dimension of *M* \**M*, where various rows correspond to different directed pairings of regions – that is, both the pair (*i, j*) and the pair (*j, i*) were included – and only linkages relating to self-loops – that is, of the type, i.e., (*i, i*) were eliminated from consideration [38]. The resulting representation of MC matrices was large since:

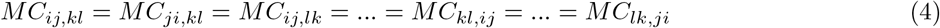

MC matrices captured higher level correlation between triplets or quadruplets of ROIs, as compared to FC matrices.

### 2.3 Node-Edge Connectivity Based Graph Attention Network for Classification

MC matrices are a higher-order correlation feature set that tells about edge connectivity between nodes. We have proposed a generalized Ex-NEGAT model that uses an attention matrix by applying a sub-graph creation approach, which helped to reduce the execution time of information between the nodes as well as capture the representation of MC matrices. We provided MC matrices as input to the proposed model. We created a custom plugin that can parse the structure files of these MC matrices. The plugin extracted important characteristics like the computational graph topology and certain node-edge features by partitioning matrices into multiple subgraphs. This subgraph splitting approach saved computing complexity while providing a rich context for the model. The graph representation showed that nodes’ actions might dramatically affect their near neighboring nodes because of their intrinsic interconnection. However, including all neighbors for each node, particularly those at greater distances, was computationally expensive and generally not required. We propose a methodology leveraging a Depth-First Search (DFS) approach to optimize the subgraph depth parameter *d*. The reason behind using the DFS approach instead of the manual selection of the value of *d* was its ability to explore the graph by traversing down one branch as far as possible before backtracking, as well as allowing for dynamic adaptation to the local structure of the graph.

#### 2.3.1 Optimization of Subgraph Depth using DFS

To optimize the subgraph depth parameter d, we employ a Depth-First Search (DFS) algorithm to systematically explore the graph *G*(*V, E*) and select subgraphs *G′* (*V ′, E′*) for each node *n* based on a maximum depth of *d*. The subgraph *G′* is constructed to include node *n* and its neighboring nodes up to a maximum *d* depth. Here, we selected value of maximum depth *d* = 5. Formally, let *G*(*V, E*) represent the input graph, where *V* is the set of vertices and *E* is the set of edges. For each vertex *n* ∈ *V*, we define *G′* (*V ′, E′*) as the subgraph containing *n* and its neighbors within a depth limit of *d*. The vertex set *V ′* of *G′* consists of all nodes within this specified distance, while the edge set *E′* comprises all edges connecting these nodes. The process begins by iterating over each vertex *n* ∈ *V* in the graph *G*(*V, E*). The DFS algorithm explores neighboring nodes up to a maximum *d* depth at each iteration. If the distance from node *n* to a neighboring node *v* is less than or equal to *d, v* is added to the vertex set *V ′* of the subgraph *G′*. Similarly, all edges connecting *n* and its neighbors within the specified distance are added to the edge set *E′* of *G′*. By systematically traversing the graph using DFS and constructing subgraphs *G′* (*V ′, E′*) for each node *n*, we create a series of subgraphs *G*_*set*_ that capture the local structure and connectivity patterns around each node within the specified depth limit *d*. Within each subgraph, the attention matrix is applied to assign weights to the edges based on their relevance, which is learned during the training phase. This allows the Ex-NEGAT model to focus on the most significant connections, enhancing the interpretability and performance of the network. The attention mechanism ensures that the most informative relationships are given priority, thus reducing noise from less relevant nodes and edges.

#### 2.3.2 Model-Architecture

The traditional Graph neural networks were considered only node features. In the current work, we proposed an attention mechanism for each edge that could consider their multidimensionality and complexity. MC matrices were higher-order correlation feature set that gave correlation values for each edge. We hypothesized that the correlation reflected by edge weight might not represent the attention required for connected nodes. To test our hypothesis, we applied a linear transformation to both node feature set *h*_*i*_ and edge feature *e*_*ij*_ set that extracted high-level associated feature set by generating a new feature representation:

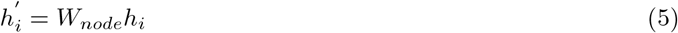

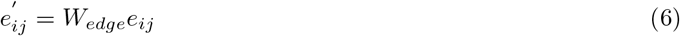

Where *W*_*node*_ and *W*_*edge*_ were indicating weight matrices. However, transformed node and edge features were used to calculate separate attention coefficients, where the attention coefficient for node *i* was computed using equation 7:

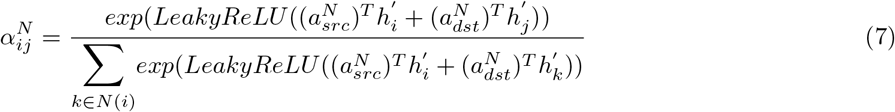

where 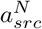 and 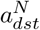 were representing learnable parameters for source and destination and *N* (*i*) = neighbours of node *i*. Similarly, calculated attention coefficient for edge *e*_*ij*_ using equation 10:

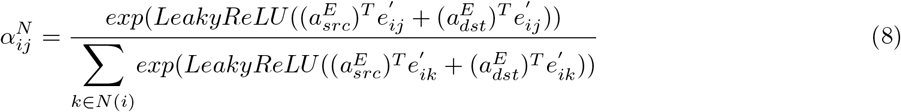

where 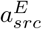 and 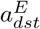 were representing learnable parameters for edge features. Finally, we updated node and edge feature weight according to attention coefficients:

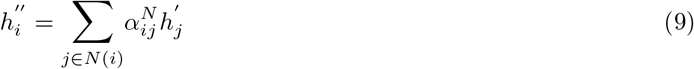

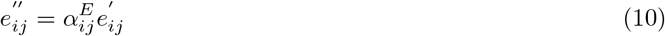

Lastly, we concatenated node and edge features and applied a linear layer to reduce dimensionality:

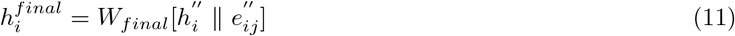

Where *W*_*final*_ = Weight matrix and 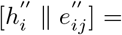 Updated node and edge features.

Finally, we used Multi-layer perception (MLP) to make predictions for individual graphs of ASD and TD based on the final feature map 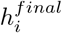. The MLP layers involved two linear layers with dropout = 0.2 and ReLU function following first linear layer to improve robustness of the proposed model.

#### 2.3.3 Training and Testing Process

We trained our model in different ways, in which the learning rate was set to 0.0001, the batch size was 64, and the number of epochs was set to 100. The L2 regularization parameter has been set to 0.0001 for all the linear layers to prevent overfitting. We utilized the Adam optimizer, and cross-entropy loss is as loss function. In addition, to prevent the issue of the model overfitting, we use a technique called label smoothing, as illustrated in the equation (12) [40]:

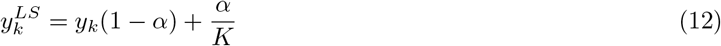

##### Algorithm 1

Pseudo-code to implement NEGAT

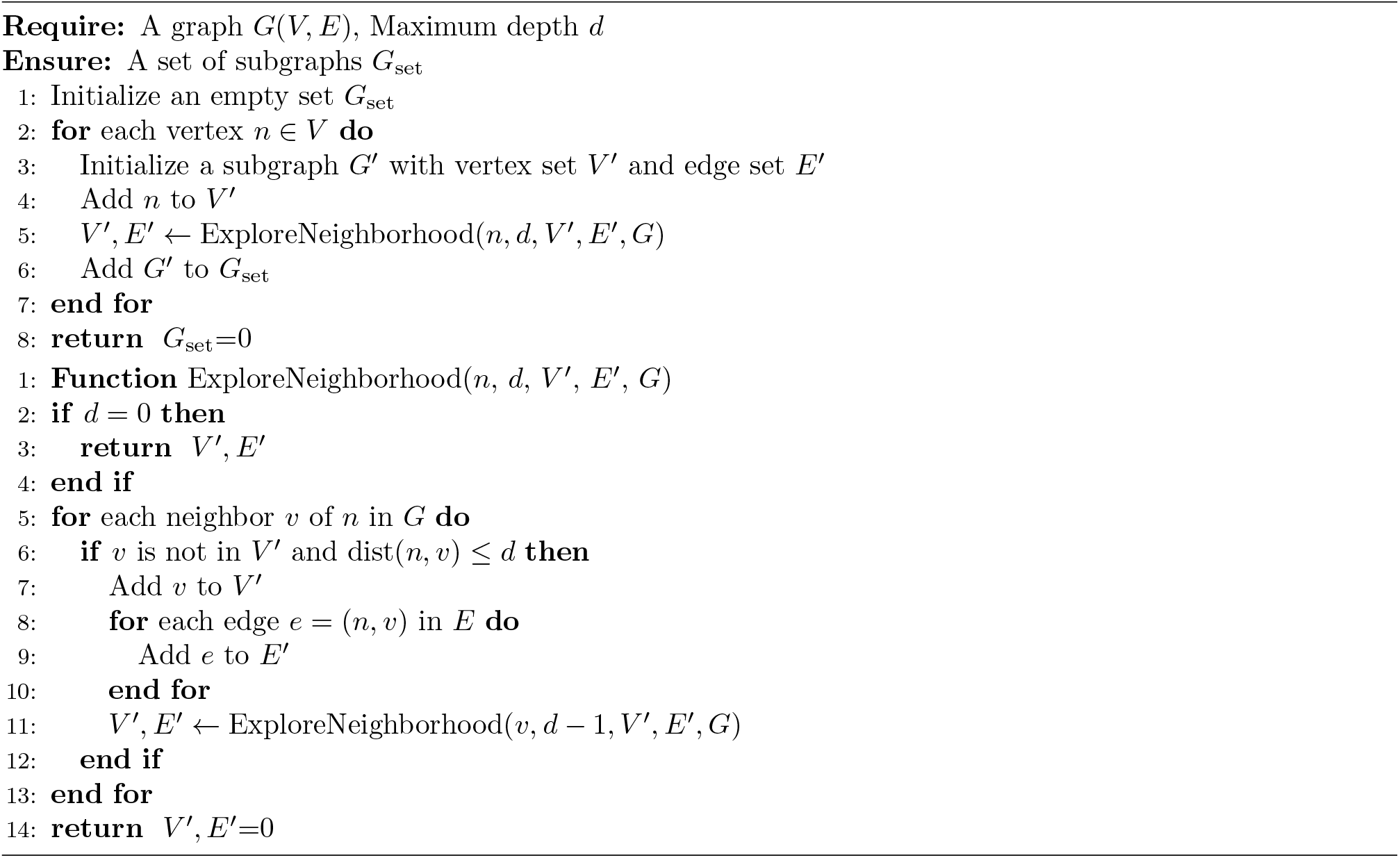

Where K = 2, that was indicating no. of class, *y*_*k*_ ∈ (0, 1) = true labels, 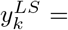 corresponding smoothing label, and *α* = 0.1. The loss function was calculated using equation (13):

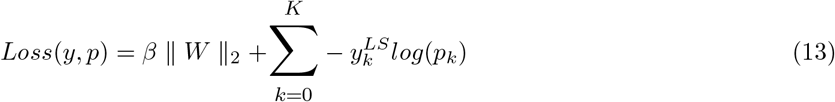

Where W = weight of linear layer, y = real label (after label smoothing), p = predicted label, *p*_*k*_ = predicted probability of class k.

#### 2.3.4 Classification Evaluation

The performance of the proposed model was tested in 3 different ways to make it more robust, explainable, and generalization: a) we trained the model on ABIDE I dataset and performed testing on ABIDE II dataset; b) we performed 5-fold cross-validation across all the subjects; c) we also used leave-one-out approach in which, we trained model on complete ABIDE I+II except one site, and perform testing on that site data as an independent dataset. We repeated this process for all sites, including ABIDE I and II datasets. We also trained different types of GNN models like graph convolutional network (GCN) [41], Chebyshev Convolution [42], Residual GCN (ResGCN) [43], GraphSAGE [44] to cross-check the proposed model’s performance using MC matrices. To validate the novelty of the proposed meta-connectivity feature set, we also performed classification using FC and dFC matrices.

### 2.4 Symptom-Severity score prediction for ASD Samples

We identified the association between neurobiological features, i.e., Meta-connectivity matrices and symptom severity scores. We investigated whether resting-state Meta-connectivity could identify ASD symptom severity scores for each individual. For this purpose, we used Connectome-based Prediction Modelling (CPM), in which each participant’s meta connectivity matrix and behavioral scores were subjected to CPM model [45]. Previously, the authors used CPM approach to behavioral scores using node-centric features like functional connectivity [45, 7]. There is no such framework, in which CPM approach is implemented using an edge-centric feature, i.e., Meta-connectivity matrices. The initial step involves splitting the input data into training and testing sets. In this study, we trained the model on ABIDE I ASD samples and performed testing on ABIDE II ASD samples. Subsequently, across all participants in the training set, each connection in the connectivity matrices is associated with the behavioral measures using various linear regression techniques, such as Pearson’s correlation, Spearman’s correlation, or robust regression. After the linear regression, the most significant connections were singled out for further analysis. Typically, prominent connections were selected through statistical significance testing, i.e., chosen connections with correlation values exceeding a predefined threshold (0.4 with p*<*0.05). For each participant, the most vital connections are consolidated into a single summary value. This is commonly done by summing the strengths of the connections. Next, a predictive model is constructed, assuming a linear relationship between the summary value of connectivity data (independent variable) and the behavioral variable (dependent variable). Subsequently, summary values are computed for each participant in the testing set, and these values are input into the predictive model.

## 3 Results

### 3.1 Computation of Meta-Connectivity Matrices

In the current study, we extracted time-series signals from ToM, DMN, CEN, and SN brain networks and calculated functional connectivity matrices for each individual. The FC matrices have a unique symmetry, and despite their large N × N size, they only have *L* = *N* (*N* − 1)/2 unique entries, representing the lower triangular part of the matrix. As the output, FC matrix appears as a *N* * *N* matrix. We also calculated dFC stream, that output in a 3D tensor sized *N* * *N* * *F*, where F was no. of frames. We chose window sizes 20 and 30 as per the literature [39]. However, each FC matrix was shown in the ‘vector’ format that outputs a vector of size *L* * 1, and dFC stream yielded a 2D matrix sized *L* * *F*, with each frame in vector format. The ‘vector’ format is advantageous because it produces more memory-efficient output, which is essential for large datasets. It also naturally represents FCs as points in a vector space, making it easier to create dimensionally reduced representations of the dFC stream using methods like t-stochastic neighborhood embedding. After that, we calculated MC matrices for each individual of size 600*600. To validate the results, we performed multiple one-sample t-tests with a p-value*<*0.01 and applied FDR correction.

### 3.2 Classification of ASD and TD Individuals using Proposed Ex-NEGAT Model

#### 3.2.1 Performance of Proposed Method using Meta-connectivity Matrices

In the current study, we proposed an attention-based graph neural network in which we divided the graph into a sub-graph upto a maximum depth *d* using the DFS approach, which helped to identify a suitable value for depth *d*. Initially, we chose the maximum value of *d* = 5, but we observed that value of *d* = 3 gave the best possible results, i.e., high accuracy with less execution time as reported in Table 4. So, we reported all the results using value of *d* = 3. To classify ASD samples from neuro-typicals, we subjected resting-state MC matrices of size 600*600 to the proposed Ex-NEGAT model. Here, we considered and performed the complete analysis using window sizes 20 and 30 sec. Firstly, we performed normal training and testing processes on ABIDE1 dataset and achieved an average accuracy of 92% with F1-score of 0.94 for window size of 20 sec and average accuracy of 91% with F1-score of 0.90 for window size of 30 sec. To make the model generalizable, we trained the model ABIDE1 dataset and performed testing on the ABIDE 2 dataset. We achieved 88% average accuracy with F1-score of 0.89 for window size of 20 sec and 83% average accuracy with F1-score of 0.85 for window size of 30 sec. We observed that we were getting better results using window size of 20 sec (Refer to Figure 4 and Table 4).

**Figure 4:**
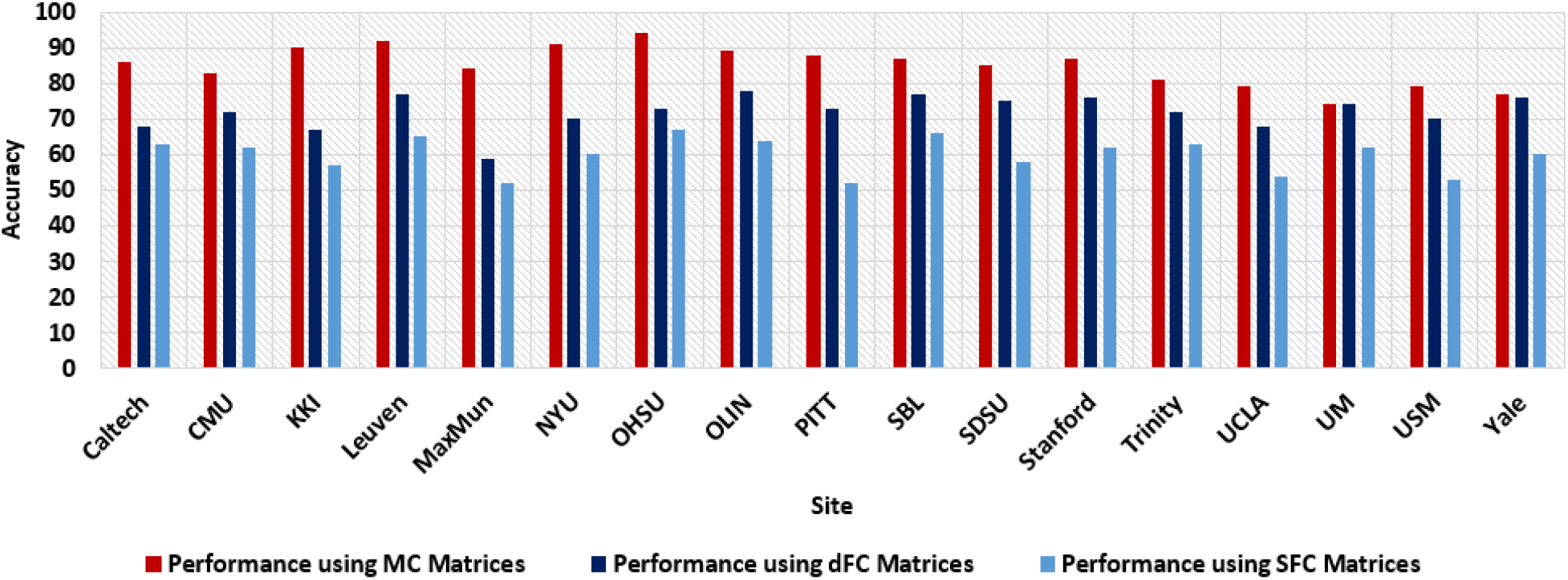
Performance of proposed model using Leave-one-out approach. The results suggested that the proposed model outperforms using the proposed meta-connectivity as a feature set.

#### 3.2.2 Comparison of Performance using Meta Connectivity and Traditional Feature sets like Static and Dynamic Functional Connectivity Matrices

An extensive comparative analysis was conducted to assess the proposed model’s performance, leveraging the novel meta-connectivity feature set against the conventional feature set. This entailed the execution of classification tasks using both static and dynamic feature sets. The Ex-NEGAT model was trained using functional connectivity (FC) matrices derived from ToM, DMN, CEN, and SN on the ABIDE I dataset, with subsequent testing on the ABIDE II dataset. We achieved an average accuracy of 63% with F1-score of 0.62 using FC matrices on unseen data. We again trained the Ex-NEGAT model using dynamic functional connectivity (dFC) of each individual on ABIDE I dataset and performed testing on the DFC of ABIDE II dataset. We achieved average accuracy of 70% with F1-score of 0.71 for window size of 30 sec and average accuracy of 74% with F1-score of 0.73 for window size of 20 sec. We were again getting better results using window size of 20 sec. There was an improvement in performance using dFC matrices as feature set compared to FC as feature set (Refer to Table 4).

These findings collectively underscore the limitations of conventional feature sets in effectively classifying the ASD samples when confronted with unseen data. In contrast, the MC matrices as a feature set, which emphasizes higher-order correlation capturing edge strength variations over time for each individual, exhibited remarkable performance. It successfully distinguished the ASD population in the context of specific brain networks when applied to unseen data. This underscores the efficacy of the proposed framework in leveraging MC matrices as a feature set for specific brain networks, illuminating a promising avenue for further exploration.

#### 3.2.3 Cross-validation using Five-fold Cross-validation and Leave-one-out Approaches

The classification framework’s evaluation involves utilizing two distinct approaches to mitigate overfitting and ensure a robust and generalized assessment of its classification performance. The initial approach entails implementing five-fold cross-validation across all subjects, with a specific value of k (in this study, k =5). The second approach adopts a more stringent method known as the Leave-one-out approach. Under this strategy, one particular site’s data is designated as an independent testing dataset, while the data from all other sites collectively constitute the training dataset. Using five-fold cross-validation, we achieved average accuracy of 81% with F1-score of 0.78 for window size of 30 sec and average accuracy of 82% with F1-score of 0.83 for window size of 20 sec. While using the Leave-one-out method, we achieved average accuracy of 78% with F1-score of 0.79 for window size of 30 sec and average accuracy of 85% with F1-score of 0.84 for window size of 20 sec. We observed that leave-on-out gave better-generalized results in which each site data was treated as unseen data for testing (refer to Table 6 and Figure 4).

### 3.3 Prediction of Symptom Severity Score

To identify robustness of MC matrices as higher order correlation feature set, we performed a prediction of symptom severity scores for each ASD sample using the CPM approach. We used MC matrices of ABIDE I dataset for training the model and performed testing on unseen data, i.e., MC matrices of ABIDE II dataset. Our proposed model was able to give an average accuracy of 85% with an F1-score of 0.86. We also performed validation using 5-fold cross-validation approach. Our model predicted ADOS-MODULE, ADOS-STEREO, ADOS-COMMUNICATION, FIQ, and ADOS-TOTAL scores more accurately, whereas it did not give accurate results for ADOS-SOCIAL scores. We also performed prediction of symptom severity scores using FC matrices. Using FC matrices, we got an average accuracy of 75% with an F1-score of 0.76. Our approach is novel as no framework in the literature identified higher order correlation (MC matrices) as a predictive measure for the prediction of symptom severity scores (refer to Figure 5). Even in large testing data, the proposed framework was able to predict symptom severity scores up to a considerable range.

**Figure 5:**
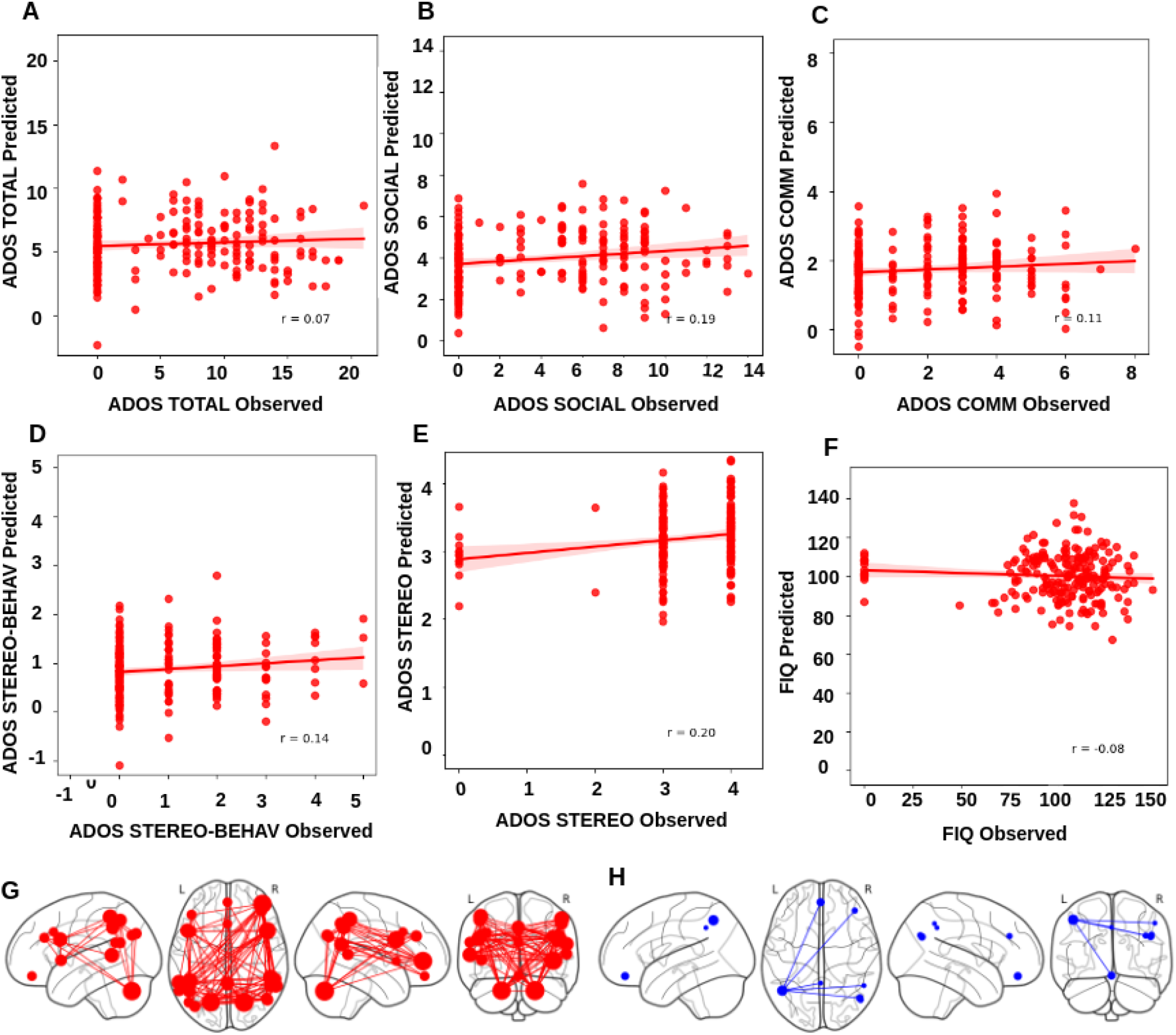
Prediction of Symptom-severity scores (ADOS-Total, ADOS-Social, ADOS-Module, ADOS-Communication, ADOS-Stereo, and FIQ scores) using Connectome-based prediction modeling in which Meta-connectivity was used as feature set. **A)** shows results for brain regions whose edge connectivity contributed positively to the prediction. Whereas **B)** depicts results for brain regions whose edge connectivity contributed negatively to the prediction.

## 4 Discussion

We developed a novel explainable and generalized Node-edge connectivity-based graph attention neural network (Ex-NEGAT) model which could capture complex patterns exhibited by dynamic functional connectivity in the resting state during atypical neurodevelopment and is considered as a correlate of cognitive processing. In this work, we make a substantial gain and acquire an important insight about time varying nature of brain networks during early developmental changes by conceptualizing dFC as a flow across morphing connectivity configurations, our notion of dFC speed quantifies the rate at which FC networks evolve in time which serves as an important imaging biomarker for accurate classification between ASD and TD and predicting symptom severity scores at the individual subject level. Here, we probe the hypothesis that variations of resting state dFC flow characterized by meta-connectivity are selectively interrelated within specific functional subnetworks (ToM, DMN, CEN, and SN networks) and associated with deficits in social cognition, communication, and interaction abilities frequently reported in the extant literature [1, 2].

### 4.1 Advantages of using meta-connectivity feature to discover new ASD biomarkers

A growing body of recent works including previous works from our group indicates that aberrations in Autism Spectrum Disorder (ASD) are frequently associated with functional connectivity (FC), dynamic functional connectivity (dFC) between salient brain regions [46, 47, 39, 38]. Many similar studies using feature selection based on graph data has been carried out for other mental disorders such as Schizophrenia and Attention Deficit Hyperactivity Disorder(ADHD) [48, 49, 50]. According to the existing literature, large-scale brain networks anchored in ToM, DMN, CEN, and SN networks and their connectivity profiles based on structural and functional neuroimaging to index atypical development in autistic individuals and subserve as crucial biomarker of brain graphs [51, 52, 53, 54]. However, to our knowledge, the brain’s functional network constitutes a complex spatio-temporal network structure which entails both connectivity and dynamics. Dynamics during connectivity switching driven by various internal states importantly attributes neural flexibility and the mutual influence observed between graph-edges over time or time-dependent functional links [55, 56, 57]. Hence, it is highly probable that whole brain meta-connectivity between these subnetworks may also exhibit mutual influences, a notion that has very rarely been addressed in research. Identifying new biomarkers related to diseases holds significant importance for diagnosis and treatment. Thus, analyzing the interactions between meta-connectivity which describe resting state dFC as a smooth flow across continually morphing connectivity configurations based on characterizing dFC streams and their relationship with mental disorders may offer a novel perspective for discovering biomarkers(refer to Table 2).

### 4.2 Advantages of using MC matrix and attention-based GNN

fMRI data possess both temporal and spatial properties, many recent works now analyze and process data based on their temporal and spatial features separately. One of the frequently employed methods to apply traditional machine learning algorithms or artificial neural networks(ANN) to graph data is to ignore the structure of the graph and treat the input edge weights as a vector of features [58]. However, the caveat with many of these approaches are that they ignore the crucial topological and statistical relationship among functional brain modules and large-scale brain networks which is important for the emergent properties of brain functions and constraining dynamics [59]. Another recent model has introduced a Spatial-Temporal Attention Graph Convolutional Network (STAGCN) for the classification of functional connectivity [60]. In the spatial domain, this model utilizes attention-enhanced graph convolutional networks to process the topological features of brain regions. In the temporal domain, it employs a multi-head self-attention method to capture the temporal relationships between different dynamic Functional Connectivity (dFC) [60, 23]. Although, many recent attempts leverages the temporal and spatial characteristics of fMRI data, the accuracy of ASD diagnosis leaves something more to be desired and fell well short of the expected classification accuracy [60, 61]. An alternative approach could be to treat adjacency matrix of the graph as an image and extract features using traditional CNN models. However, as has been shown the spatial proximity in the adjacency matrix elements does not necessarily always correspond with topological locality of the graph data [59, 62]. However, in this work we differed from all the previous approach and used a node-edge centric spatio-temporal meta-connectivity feature to train a novel generalized Ex-NEGAT model. This MC meta-module may be at certain times coordinated (when the co-fluctuating links are in a large strength transient) and thus form a *FC*(*t*) module in the conventional sense. This link sets that form different meta-modules will fluctuate toward large or weak strengths at different independent times. Importantly, this transient property of the links and their cofluctuations at different time windows seem to be a key feature that plays an important role in the diagnosis task of brain diseases as demonstrated in this work 4 and Table 4). Compared with the previously employed methods, we create an attention-based network by applying a sub-graph creation approach reducing computational time and interpretability of our model based on meta-links between relevant brain networks. Our attention-based GNN make the classification model more transparent and enhancing the possibility for detecting potential spatio-temporal imaging biomarkers of ASD. Moreover, instead of using convolution based method used earlier on FC by [61] the subgraph creation approach of GNN can aggregate spatio-temporal information and extract node-edge centric features on the non-Euclidean graph data according to graph topological properties [63]. It can generate more subgraph specific information based on meta-modules that is beneficial to the Autism classification task. In the next subsection, we discuss in details about how this approach also significantly reduces computational time.

### 4.3 Advantages of using MC subgraphs to reduce computational time

To classify between ASD and TD samples, we employed meta-connectivity matrices as higher-order correlation features. Additionally, static and dynamic FC measures were also calculated for each individual subjects to compare the performance of all three connectivity features in accurately classifying ASD and TD. Our findings revealed hypo-connectivity within edges for the ASD group and on the contrary, hyper-connectivity within edges for the TD group (see Table 7). Meta-connectivity matrices from the ABIDE I dataset were utilized for training the Ex-NEGAT model, with testing conducted on the ABIDE II dataset. Further, our proposed generalized Ex-NEGAT model converted MC matrices into graphs and then divided them into subgraphs with maximum depth *d* using the DFS approach. In order to determine the optimal depth parameter in DFS, denoted as *d*, we initially explored up to *d* = 5, corresponding to traversing sub-graphs up to the fifth-order neighbor. However, increasing *d* beyond 5 escalated execution time without substantial improvement. Traversing to the first and second-order neighbors (*d* = 1 or 2) yielded less accurate predictions but reduced execution time. Optimal prediction performance was observed at *d* = 3, with reasonable execution time. Further exploration of the fourth and fifth-order neighbors showed minimal improvement in prediction accuracy at the cost of increased execution time. Our analysis using Depth-First Search (DFS) determined that traversing up to the third-order neighbor (*d* = 3) offered the best model performance (see Table 4). The proposed framework is unique for the following reasons: a) Use of a novel feature set that has not been employed in classification, b) unique attention-based GNN architecture based on subgraph creation, and c) the proposed model could reduce the execution time of information from one node to another, which could help in reducing computational resources and time-complexity. To make our model generalizable, training was performed on ABIDE I dataset and performed testing on ABIDE II dataset (Refer to Table 3).

**Table 3:**
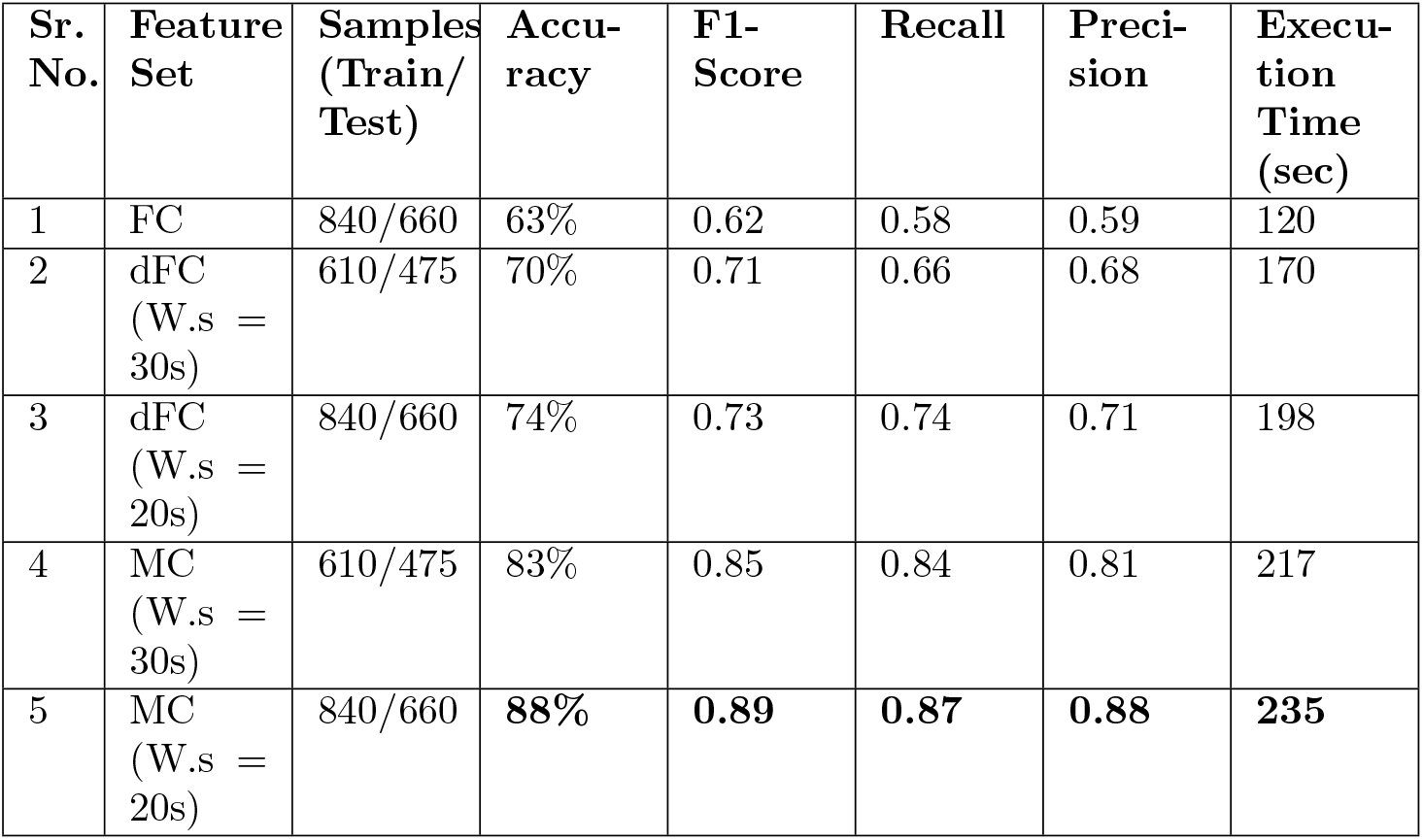
Performance of proposed Ex-NEGAT model using FC,dFC, and MC matrices as feature set with *d* = 3, where W.s represents window size.

| Sr. No. | Feature Set | Samples (Train/Test) | Accuracy | F1-Score | Recall | Precision | Execution Time (sec) |
| --- | --- | --- | --- | --- | --- | --- | --- |
| 1 | FC | 840/660 | 63% | 0.62 | 0.58 | 0.59 | 120 |
| 2 | dFC (W.s = 30s) | 610/475 | 70% | 0.71 | 0.66 | 0.68 | 170 |
| 3 | dFC (W.s = 20s) | 840/660 | 74% | 0.73 | 0.74 | 0.71 | 198 |
| 4 | MC (W.s = 30s) | 610/475 | 83% | 0.85 | 0.84 | 0.81 | 217 |
| 5 | MC (W.s = 20s) | 840/660 | **88%** | **0.89** | **0.87** | **0.88** | **235** |

**Table 4:** Performance of proposed model using different values of depth *d*. The table is highlighting Accuracy, F1-Score, Precision, Recall, and Execution time in seconds.

| Value of $d$ | Performance using FC Matrices | Performance using dFC Matrices | Performance using MC Matrices |
| --- | --- | --- | --- |
| - | Acc.(%), F1-s, Prec., Rec., Exe.(s) | Acc.(%), F1-s, Prec., Rec., Exe. (s) | Acc.(%), F1-s, Prec., Rec., Exe. (s) |
| $d=1$ | 54%, 0.53, 0.54, 0.51, 80 | 65%, 0.67, 0.63, 0.64, 120 | 71%, 0.73, 0.68, 0.67, 156 |
| $d=2$ | 59%, 0.61, 0.57, 0.59, 103 | 69%, 0.67, 0.68, 0.66, 159 | 76%, 0.77, 0.71, 0.72, 197 |
| $d=3$ | **63%, 0.62, 0.58, 0.59, 120** | **74%, 0.73, 0.74, 0.71, 198** | **88%, 0.89, 0.87, 0.88, 235** |
| $d=4$ | 66%, 0.67, 0.63, 0.65, 135 | 73%, 0.76, 0.72, 0.70, 220 | 89%, 0.87, 0.88, 0.85, 329 |
| $d=5$ | 65%, 0.65, 0.63, 0.66, 176 | 75%, 0.73, 0.76, 0.71, 308 | 87%, 0.85, 0.83, 0.81, 433 |

**Table 5:** Performance comparison of multiple models including proposed model in the classification of ASD from neuro-typical samples using MC matrices as feature set.

| Sr. No. | Classifier | Accuracy | F1-Score | Recall | Precision | AUC |
| --- | --- | --- | --- | --- | --- | --- |
| 1 | SVM | 50% | 0.48 | 0.46 | 0.45 | 0.52 |
| 2 | Random forest | 56% | 0.55 | 0.53 | 0.54 | 0.55 |
| 3 | LSTM-RNN | 71% | 0.72 | 0.69 | 0.70 | 0.69 |
| 4 | 2d-CNN | 75% | 0.74 | 0.73 | 0.72 | 0.76 |
| 5 | CapsNet | 78% | 0.78 | 0.76 | 0.770 | 0.76 |
| 6 | Graph convolutional network (GCN) | 71% | 0.69 | 0.67 | 0.70 | 0.73 |
| 7 | Chebyshev Convolution (GCNN) | 75% | 0.74 | 0.73 | 0.75 | 0.76 |
| 8 | Residual GCN (Res-GCN) | 76% | 0.77 | 0.75 | 0.76 | 0.73 |
| 9 | Graph SAGE | 80% | 0.81 | 0.78 | 0.77 | 0.82 |
| 10 | Proposed Ex-NEGAT | **88%** | **0.89** | **0.87** | **0.88** | **0.86** |

**Table 6:** Validation of performance of the proposed EX-NAGET model using the Leave-one-out approach.

| Site | No. of Samples | No. of M/F | Performance using MC Matrices | Performance using dFC Matrices | Performance using FC Matrices |
| --- | --- | --- | --- | --- | --- |
| - | - | - | Accuracy (%), F1-score | Accuracy (%), F1-score | Accuracy (%), F1-score |
| Caltech | 43 | 29/14 | 86%, 0.84 | 68%, 0.69 | 63%, 0.62 |
| CMU | 37 | 31/6 | 83%, 0.85 | 72%, 0.73 | 62%, 0.61 |
| KKI | 65 | 46/19 | 90%, 0.91 | 67%, 0.66 | 57%, 0.59 |
| Leuven | 85 | 65/20 | 92%, 0.90 | 77%, 0.78 | 65%, 0.67 |
| MaxMun | 72 | 64/8 | 84%, 0.85 | 59%, 0.60 | 52%, 0.53 |
| NYU | 312 | 240/72 | 91%, 0.89 | 70%, 0.72 | 60%, 0.61 |
| OHSU | 39 | 39/0 | 94%, 0.92 | 73%, 0.73 | 67%, 0.68 |
| OLIN | 48 | 38/10 | 89%, 0.88 | 78%, 0.77 | 64%, 0.65 |
| PITT | 76 | 63/13 | 88%, 0.90 | 73%, 0.73 | 52%, 0.53 |
| SBL | 40 | 40/0 | 87%, 0.84 | 77%, 0.73 | 66%, 0.68 |
| SDSU | 45 | 34/11 | 85%, 0.87 | 75%, 0.72 | 58%, 0.55 |
| Stanford | 47 | 35/12 | 87%, 0.86 | 76%, 0.73 | 62%, 0.64 |
| Trinity | 71 | 71/0 | 81%, 0.83 | 72%, 0.74 | 63%, 0.60 |
| UCLA | 138 | 113/25 | 79%, 0.82 | 68%, 0.67 | 54%, 0.55 |
| UM | 211 | 173/38 | 74%, 0.77 | 74%, 0.73 | 62%, 0.63 |
| USM | 94 | 94/0 | 79%, 0.78 | 70%, 0.72 | 53%, 0.54 |
| Yale | 77 | 55/22 | 77%, 0.79 | 76%, 0.76 | 60%, 0.63 |

**Table 7:**
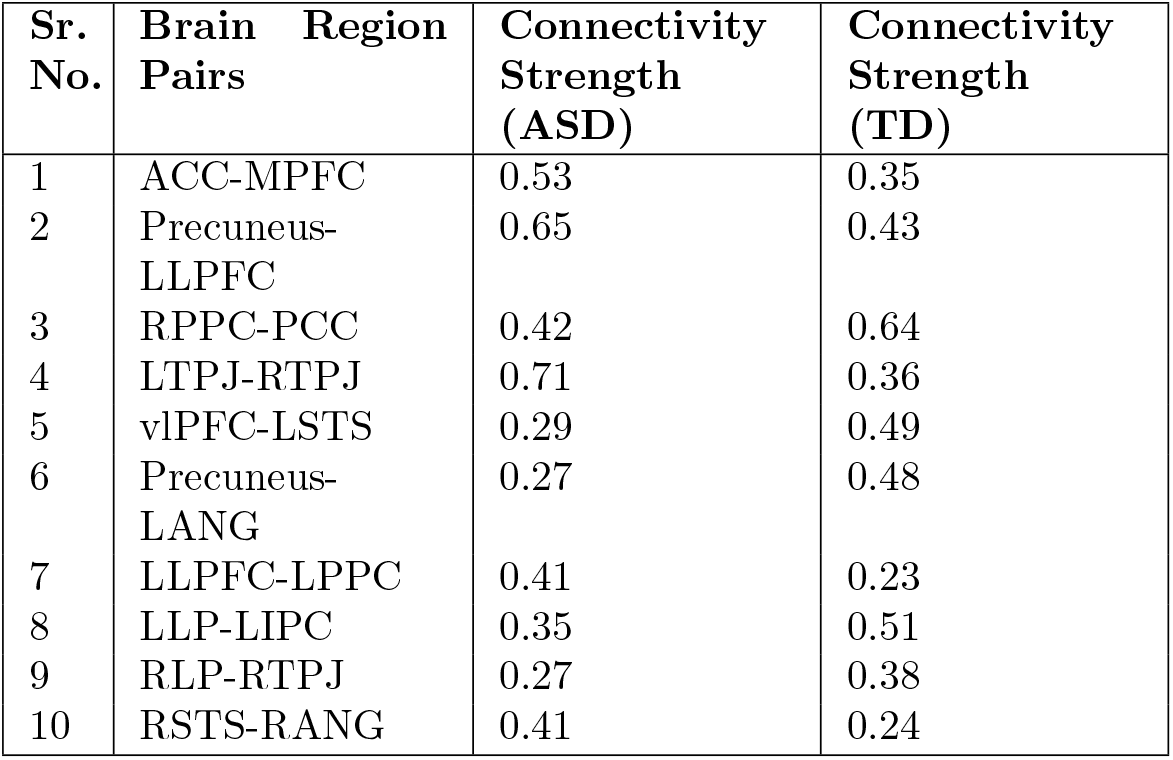
Table shows the top 10 pairs of nodes whose edge connectivity contributed most to the prediction. The table shows edge connectivity strength for ASD and TD groups.

### 4.4 Predicting symptom severity in ASD using MC matrices and identifying relevant ROIs

To check robustness of MC matrices as higher-order correlation feature set, we performed prediction of symptom severity scores using the CPM approach in which meta-connectivity matrices were subjected as input. Our approach notably demonstrated that resting-state meta-connectivity between ToM, DMN, CEN, and SN sub-networks could classify ASD samples from TD samples accurately on unseen datasets, as well as this feature set could also predict symptom severity scores without requiring any additional feature-engineering approach (Refer to Figure 1). In the previous works, [64, 65], the authors tried to predict symptom severity using functional connectivity between ROIs which relies on spatial features of fMRI data, but there is no proposed framework based on attention-based GNN that predicted symptom severity and diagnostic scores using meta-connectivity spatio-temporal features in functional neuroimaging. We trained a CPM model based on MC matrices derived based on ABIDE I ASD samples and performed testing on ABIDE II samples. Our model accurately predicted ADOS-Total, ADOS-Social, ADOS-Module, ADOS-Communication, ADOS-Stereo, and FIQ scores. We also identified ROIs whose meta-connectivity edges contributed positively and negatively to the symptom severity score prediction. We found that edges between Precuneus, Medial prefrontal Cortex (MPFC), Left Superior Temporal Sulcus (LSTS), Right Lateral Prefrontal Cortex (RLPC), Right Superior Temporal Sulcus (RSTS), Right Posterior Parietal Cortex (RPPC) were contributed as negatively in prediction, whereas all other edges between other ROIs contributed positively(Refer to Figures 4). Extant literature suggests that a critical social and cognitive dysfunction in ASD is the impaired ability to decode the mental states such as beliefs, emotions and intentions of self as well as others, and altered DMN may be an important neurobiological feature of these deficits [66, 67]. Previous neuroimaging work has consistently suggested that the DMN is one of the most aberrant functional networks in ASD [68, 69]. Studies of intrinsic FC in ASD reported disrupted within-network connectivity between core DMN nodes [70, 71, 39, 72, 38]. To validate the result, we also tried to predict the symptom severity scores at the individual level using DMN, ToM region specific functional connectivity, but this resulted in fairly lower accuracy on unseen data3 and 4.

The proposed explainable and generalized attention-based GNN framework and feature set not only distinguish ASD from TD but also predicted their symptom severity scores on large unseen data. We observed that the SN contributed the most to the classification and prediction of ASD. This is consistent with existing literature suggesting the social-brain circuit dominantly includes classic limbic areas, ventral and medial aspects of the prefrontal cortex, the anterior temporal lobes, the posterior cingulate cortex (PCC), the PCUN, the posterior temporal regions, the temporo-parietal junction (TPJ), the left IFG involved in social communication, somatosensory and anterior insular cortex [73, 4, 74, 75, 76, 60, 23], which was implicated in the neurological mechanisms in ASD. The proposed framework also reduces computational time and is a lightweight approach in contrast with whole brain approach by focusing on selective set of brain networks and node-edge centric features and consequently, minimal computational resource is necessary to run the proposed framework.

## 5 Conclusion

In summary, our discovery of robust, individualized functional brain meta-connectivity links is a promising biomarker within relevant brain regions related to social cognition in ASD diagnosis, and tracking transient dynamics of meta-connectivity in pervasive neurodevelopmental disorders. Beyond that, our innovative approach offers Ex-attention-based generalized GNN framework based on subgraphs to explore the reliable and interpretable neurobiological features from medical imaging data providing crucial insights into their clinical symptoms and advancing precision neuroimaging in brain disorders.

## Data Availability

https://fcon_1000.projects.nitrc.org/indi/abide/

